# Efficacy of Sofosbuvir, Daclatasvir and Ribavirin combination therapy in treatment naïve hepatitis C patients at tertiary care hospital of South Punjab

**DOI:** 10.1101/2021.11.23.21266653

**Authors:** Qasim Umar, Muhammad Asif Gul, Farooq Mohyud Din Chaudhary, Shehryar Kanju, Rizwan Hameed, Dure Shahwar

**Affiliations:** Nishtar Medical University & Hospital, Multan; PHD, Department of Pharmaceutics, Bahauddin Zakariya University, Multan

**Author notes:** Corresponding Author: Dr Qasim Umar, Email Address, Contact number: 03336739590, Adrress: House No. 1, Wasti Stree, Near Chungi No. 6, Bosan Road, Multan. **Funding:** No funding was received for the publication of this article.

**Keywords:** sofosbuvir, daclatasvir, efficacy, sustained virological response, hepatitis C

## Abstract

**Introduction:** Hepatitis C has gradually become endemic in Pakistan, with infectivity rates one of the highest in the world. The emergence of direct acting antivirals (DAAs) has become a ray of sunshine in eradicating this menace from this region. The combination of sofosbuvir, daclatasvir and ribavirin (SOF/DACLA/RIBA) has had phenomenal success all over the world in eradicating this virus. Our study aims to see the effectiveness of this regime in this part of the world.

**Methods:** After approval from the institutional review board (IRB), retrospective analysis of data of treatment naïve patients who have been treated with the above mentioned regimen was collected to assess the efficacy by calculating the sustained virological response (SVR) at 12 weeks after completion of therapy.

**Results:** Data of 300 patients (172 females, 128 males) was collected. Mean age was 39.66 years. Majority (almost 90%) of patients were from District Multan Age range was from 18 years to 60 years. Eighty-three percent of the patients were non-cirrhotics, 15.7% had compensated cirrhosis, while only 1 % had decompensated cirrhosis. Out of the 300 patients, 291 patients had undetectable HCV RNA on polymerase chain reaction (PCR) at 12 weeks after completion of treatment, achieving SVR rates of 97%. There was no significant association of SVR rates with gender and age of patients.

**Conclusion:** The combination of SOF/DACA/RIBA is highly efficacious for treatment of hepatitis C patients.

## Introduction

Viral hepatitis is a global health problem with hepatitis C being the most common cause. It affects almost 71 million people worldwide with almost three hundred thousand deaths recorded in 2019 according to World Health Organization [1]. Hepatitis C prevalence rate was estimated at 5% in Pakistan, with over 8 million people infected, according to National survey conducted in 2008 [2]. The prevalence in Punjab rose from 6.7% in 2008 [2] to 8.9%(14 million population) according to Bureau of Statistics, Punjab in 2017-18. Pakistan has the highest prevalence of Hepatitis C in the world [3], followed by Egypt. If left untreated it can lead to cirrhosis and its related complications, hepatocellular carcinoma and ultimately death [4-7].

For many years, the mainstay of treatment of Hepatitis C was interferon therapy with a response rate of 40-60% and high incidence of very serious side effects. Treatment was cumbersome and expensive as well. With the advent of Direct acting anti-viral drugs (DAAs), there is a paradigm shift with high success rate and very few side effects [8,9].

Over the years, significant development has taken place in manufacturing of DAAs. The Punjab Hepatitis Control program is providing free of cost treatment for Hepatitis C. Even though newer drugs are available in Pakistan, in the government hospitals sofosbuvir, daclatasvir and ribavirin combination therapy is given to patients. In this region of Pakistan, real world data regarding efficacy of this regime is scant. The aim of our study was to assess the efficacy of this regime in the eradication of this deadly virus.

## Methods

This retrospective study was conducted at the gastroenterology department, Nishtar Hospital Multan. Study was approved by the Institutional review Board of Nishtar Medical University Multan (Reference No. 18675). From the record register of Hepatitis Clinic, data of patients who came for Hepatitis C treatment in 2018 was collected. All patients who presented to the Hepatitis Clinic in 2018 with positive antibodies to HCV and detectable HCV RNA on polymerase chain reaction (PCR) were included in the study. Patients less than 18 years of age, more than 60 years of age, patients with Hepatitis B surface antigen positivity, co-morbidities such as diabetes and hypertension and any type of malignancies were excluded from the study. Three hundred patients who fulfilled the above criteria were enrolled in our study. These patients were given triple regimen for hepatitis C eradication; sofosbuvir 400mg daily, daclatasvir 60mg daily and weight-based ribavirin (1000mg in patients less than 75kg and 1200mg in patients more than 75kg weight). Confidentiality of the participants was ensured.

From the records patient’s age, gender, district of residence, stage of disease and efficacy of treatment was recorded. Disease staging was done as follows: Chronic Hepatitis C-detectable HCV RNA on PCR, normal liver and spleen on ultrasound abdomen, normal bilirubin, albumin and prothrombin time; Compensated Cirrhosis-detectable HCV RNA on PCR, coarse echogenic liver and enlarged spleen on ultrasound, Child Class A; Decompensated Cirrhosis-detectable HCV RNA on PCR, shrunken liver, enlarged spleen and ascites on ultrasound abdomen, Child Class B or C. Child Classification was according to Child Turcotte Pugh Scoring [reference]. Efficacy was assessed by undetectable HCV RNA on PCR at 12 weeks after completion of treatment.

All the data was entered and analyzed in the Statistical Package for social Sciences (SPSS) version 23 (IBM Inc). Frequencies were calculated for numerical variables while table and chi square tests were applied for categorical variables.

## Results

A total of 300 patients were included in our study. There were 172 females (57.3%) and 128 male (42.7%) patients in our study. Mean age of the patients was 39.66 years with a standard deviation of 11.2 years and age range of 18 to 60 years. Table 1 shows the district wise distribution of patients of our study. Almost 90% of the patients belonged to district Multan. Table 2 shows the distribution of hepatitis C patients according to their disease severity. More than 80 percent of patients belonged to non-cirrhotic chronic hepatitis C category.

**Table 1:**
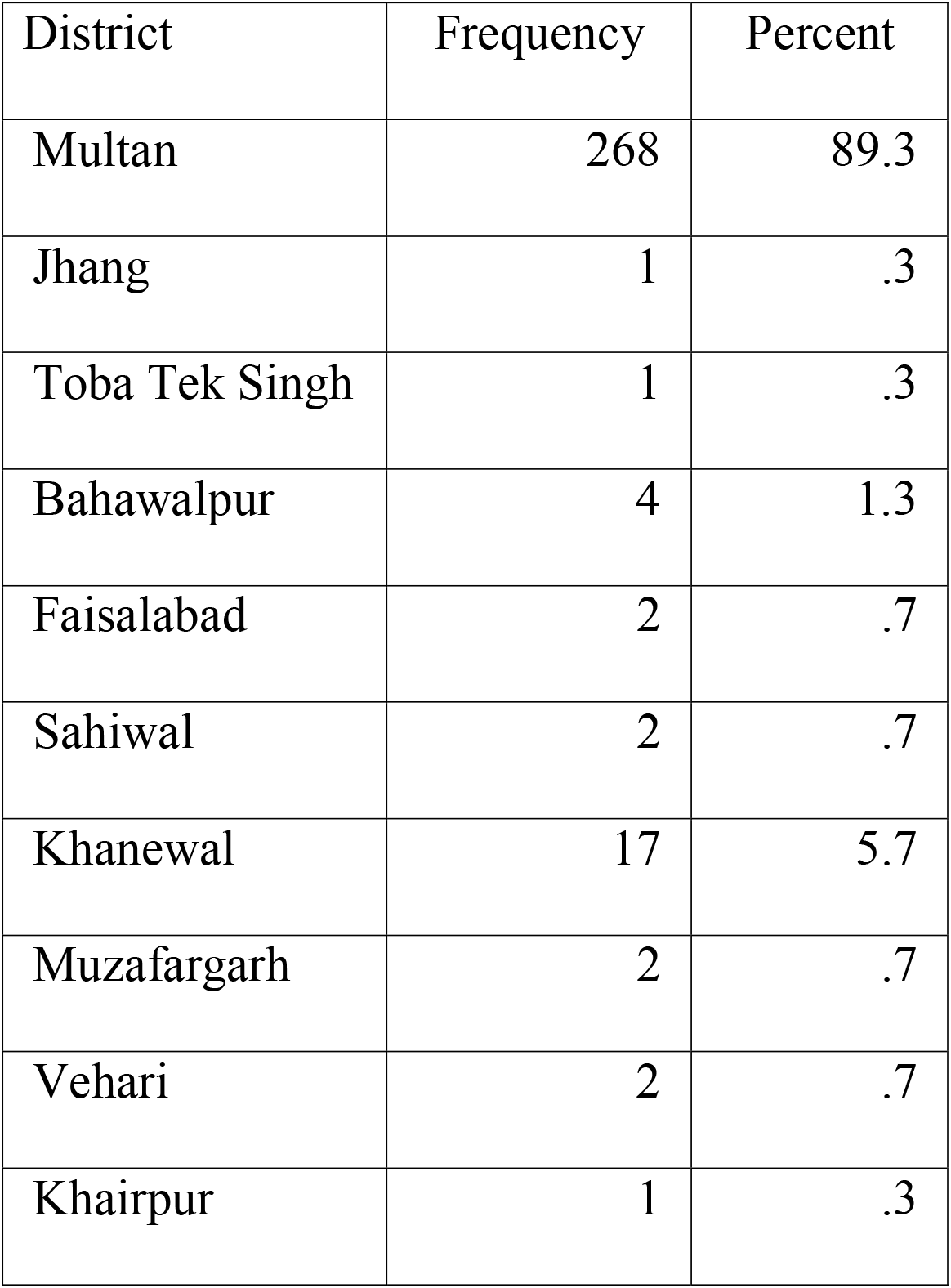
District wise distribution of patients

| District | Frequency | Percent |
| --- | --- | --- |
| Multan | 268 | 89.3 |
| Jhang | 1 | .3 |
| Toba Tek Singh | 1 | .3 |
| Bahawalpur | 4 | 1.3 |
| Faisalabad | 2 | .7 |
| Sahiwal | 2 | .7 |
| Khanewal | 17 | 5.7 |
| Muzafargarh | 2 | .7 |
| Vehari | 2 | .7 |
| Khairpur | 1 | .3 |

**Table 2:** Disease Severity of Hepatitis C patients

| Stage | Frequency | Percent |
| --- | --- | --- |
| Chronic Hepatitis | 250 | 83.3 |
| Compensated Cirrhosis<br>(CTP-A) | 47 | 15.7 |
| Decompensated Cirrhosis<br>(CTP-B/C) | 3 | 1.0 |

CTP: Child Turcotte Pugh classification; CTP-A: score less than 7; CTP-B: score 8-9; CTP-C: score 10 and above

Table 3 shows the response rate of the treatment regimen. The sustained virological response (SVR) at 12 weeks after completion of therapy was 97%. Table 4 shows the association of gender and age with the SVR. Results shows no significant difference with regards to gender or young or old age on SVR rates (p value > 0.05).

**Table 3:** Response of treatment at 12 weeks after completion of therapy

|  | Frequency | Percent |
| --- | --- | --- |
| SVR achieved | 291 | 97.0 |
| SVR not achieved | 9 | 3.0 |

SVR: Sustained Virological Response

**Table 4:** Association of age and gender with SVR

| Variable |  | SVR status at 12 weeks |  | P value |
| --- | --- | --- | --- | --- |
|  |  | SVR achieved | SVR not achieved |  |
|  |  | N (%) | N (%) |  |
| Age | < 40 years | 160 (97) | 5 (3) | 1.0 |
|  | >40 years | 131 (97) | 4 (3) |  |
| Gender | Male | 123 (96.1) | 5 (3.9) | 0.5 |
|  | Female | 168 (97.7) | 4 (2.3) |  |

SVR: Sustained Virological Response

## Discussion

The published data reveals that hepatitis c virus is endemic in Pakistan. Prevalence of Pakistani population is estimated at 6.8%. In recent years there is almost a 40% rise of sero-prevalence of hepatitis c [10]. Sofosbuvir based regimes are treatment of choice nowadays. Once daily sofosbuvir with daclatasvir are associated with high rates of eradication of this virus [11]. Our study is one of the few in South Punjab assessing the efficacy of this combination in eradication of hepatitis C.

Our study showed a remarkably high level of treatment response to antivirals. The SVR at 12 weeks after completion of therapy was recorded at 97%. Our results are in compliance with a another multicenter study of Pakistan which showed SVR rates of 94.4% in genotype 2 patients [12]. Similar response rates were observed in many other studies as well [13, 14, 15, 16, 17, 18]. Our study corroborates international data regarding high efficacy of this treatment regimen containing sofosbuvir and daclatasvir.

Sofosbuvir and daclatasvir combination is tolerated well by patients. Due to the retrospective nature of our study, side effects profile was not recorded in our study. However other studies have demonstrated good efficacy and safety of not only this combination [19], but also its predecessor sofosbuvir and ribavirin [20, 21]. Our study did not show any association of gender and age with treatment response. Majority of our patients were non-cirrhotics. Almost 15% had compensated cirrhosis. Our study reveals good treatment response in these subset of patients.

Treatment of hepatitis C virus is essential in halting the disease progression and preventing cirrhosis and its complications such as ascites, spontaneous bacterial peritonitis [22], encephalopathy, variceal bleeding [23, 24], renal failure and increased mortality. Due to the high prevalence rates of hepatitis C in this area [10, 25], eradication of this infection is of utmost importance.

There were a few limitations to our study. Our study was a single center study, included only treatment naïve patients and there were very few decompensated cirrhosis patients. Also the side effect profile for this regimen was not recorded in our data. Multicenter studies, including treatment experienced patients as well as patients with severe liver disease should be included in further studies to see efficacy of this regimen in this part of the world.

## Conclusion

Our study is in compliance with international and local data revealing high rates of response rates of this regimen. Considering the fact that SOF/DACLA regimen is quite affordable and Government of Punjab is providing this regime free of cost to patients in government hospital and also providing it at district level will enable us to effectively control this pandemic in foreseeable future.

## Data Availability

All data produced in the present study are available upon reasonable request to the authors

